# Parameter-efficient deep learning for pneumonia detection on chest X-rays: A comparative evaluation of explainable AI methods

**DOI:** 10.64898/2026.07.14.26358065

**Authors:** Benyamin Mahtabi, Ebrahim Nasr-Esfahani, Shokufeh Yaraghi

## Abstract

Pneumonia is a leading cause of infectious disease mortality worldwide, accounting for approximately 2.5 million deaths annually and 15% of deaths in children under five. Chest X-ray imaging remains the primary diagnostic tool, but accurate interpretation requires radiological expertise that is disproportionately concentrated in high-income settings, creating a diagnostic gap where disease burden is highest. Automated deep learning offers a scalable complement to specialist-dependent diagnosis, yet clinical adoption requires both high accuracy and transparent, interpretable reasoning. Convolutional neural networks (CNNs) have shown strong potential for pneumonia detection from chest X-rays, but two barriers impede clinical translation: the interpretability of black-box models and the computational feasibility of large architectures in resource-constrained settings. Explainable AI (XAI) methods such as Grad-CAM, Grad-CAM++, and Score-CAM address the interpretability barrier, yet systematic quantitative comparisons across multiple CNN architectures remain scarce. Furthermore, CNN architectures widely used for medical image classification carry high parameter counts that limit feasibility in resource-constrained settings, motivating architectures that achieve competitive accuracy with substantially fewer parameters. Here we propose a parameter-efficient deep learning framework for pneumonia detection based on transfer learning, evaluated across three CNN architectures representing distinct architectural families: EfficientNet-B0 with fine-tuning (proposed method), ResNet50, and DenseNet121, trained under identical conditions on the Kaggle chest X-ray dataset (5,863 images). Our method achieved 90% classification accuracy, outperforming both baselines while requiring 4.8× fewer parameters than ResNet50. To evaluate explainability, Grad-CAM, Grad-CAM++, and Score-CAM were applied across all three architectures and compared quantitatively using Intersection over Union against manually annotated lung segmentation masks, Insertion score, and Deletion score, with pairwise statistical validation via Wilcoxon signed-rank tests and Bonferroni correction. Findings show that classification accuracy and XAI explanation quality must be evaluated independently, and that the proposed parameter-efficient architecture offers a favorable trade-off for resource-constrained clinical deployment.

## 1. INTRODUCTION

Pneumonia remains a leading cause of infectious disease mortality, accounting for approximately 2.5 million deaths annually and representing 15% of all deaths in children under five [1]. The Global Burden of Disease Study 2023 estimated that lower respiratory infections caused 2.5 million deaths and 98.7 million disability-adjusted life years (DALYs), with preschool-aged children and adults over 70 years bearing the highest burden [2]. Chest X-ray (CXR) imaging is the primary diagnostic tool due to its low cost, rapid acquisition, and widespread availability [3]. However, accurate CXR interpretation requires radiological expertise that is disproportionately concentrated in high-income settings; countries in low-income regions report as few as 1.9 radiologists per million residents compared to 97.9 per million in high-income countries [4], creating a diagnostic gap where the pneumonia burden is highest [5]. Artificial intelligence systems trained on CXR data have been proposed as a scalable tool to mitigate this gap in resource-limited environments [6].

Convolutional neural networks (CNNs) have demonstrated strong potential to address this gap. Rajpurkar et al. introduced CheXNet, a 121-layer DenseNet achieving radiologist-level pneumonia detection performance on over 100,000 chest X-ray images [7]. He et al. introduced residual skip connections in ResNet, resolving the vanishing gradient problem and enabling training of very deep networks [8]. Huang et al. proposed DenseNet, where each layer receives feature maps from all preceding layers, encouraging feature reuse and reducing parameter redundancy [9]. More recently, Tan and Le introduced EfficientNet, which uses compound scaling — jointly optimizing network depth, width, and resolution via a neural architecture search-derived coefficient — achieving state-of-the-art ImageNet accuracy with substantially fewer parameters than prior architectures [10]. EfficientNet-B0’s 5.3M parameters represent a 4.8× reduction relative to ResNet50, making it a compelling candidate for deployment in resource-constrained clinical environments. Transfer learning using ImageNet-pretrained weights has been shown to substantially improve CNN performance on medical imaging tasks, even with small labeled datasets [11]. Several recent studies have demonstrated the feasibility of this approach for pneumonia and chest disease detection: Ayan et al. used ensemble CNNs with transfer learning for pediatric pneumonia diagnosis [12], while Ravi et al. proposed a multichannel EfficientNet stacking ensemble for lung disease detection [13]. Iwendi et al. and other groups have further confirmed that EfficientNet variants achieve competitive accuracy on chest X-ray classification while maintaining lower parameter counts relative to ResNet and VGG families [14]. Despite these advances, most comparative evaluations differ in training protocols, datasets, or the set of architectures evaluated, making direct comparison across studies difficult.

Despite strong classification performance, two fundamental barriers impede clinical translation of deep learning models. The first is the interpretability barrier: deep learning models operate as black boxes, producing predictions without explanations that clinicians can validate or trust [15]. Ghnemat et al. reviewed the XAI landscape in medical imaging classification, emphasizing that the black-box problem is particularly acute in clinical settings where decisions carry medicolegal consequences [29]. Tonekaboni et al. and others have argued that black-box medical AI violates the principles of evidence-based medicine, as clinicians cannot audit model inputs or reasoning pathways [30]. The second is the computational feasibility barrier: large models such as ResNet50 (25.6M parameters) carry memory and inference costs that can be prohibitive in low-resource settings and edge deployment scenarios [16]. Addressing both barriers simultaneously — through a compact architecture combined with rigorous explainability evaluation — is the central motivation of this work.

Gradient-weighted Class Activation Mapping (Grad-CAM) [17] computes gradients of the target class score with respect to final convolutional feature maps to produce spatially informative heatmaps. Grad-CAM++ [18] extends this approach using second-and third-order partial derivatives as pixel-wise weighting coefficients, improving localization for multiple or diffuse activation regions. Score-CAM [19] eliminates gradient dependency entirely, using forward-pass confidence scores of channel-masked images as channel weights. While these methods have been applied individually in prior medical imaging studies, systematic quantitative comparisons across multiple CNN architectures remain scarce. Ihongbe et al. conducted a human-centered evaluation of Grad-CAM and LIME in chest radiology, finding that while Grad-CAM showed better coherency and user trust, concerns remained about clinical usability [20]. Aasem and Javed Iqbal proposed an Ensemble-CAM approach combining multiple CAM-based explanations for thoracic disease localization, demonstrating that single-method evaluations may underestimate explanation quality variability across architectures [21]. Suara et al. raised specific concerns about whether Grad-CAM explanations in medical images are genuinely discriminative, arguing for more rigorous quantitative evaluation [22]. Erukude et al. compared ResNet50 and DenseNet121 with Grad-CAM on chest X-rays but evaluated only two architectures and one XAI method without quantitative metrics [24], and The Wilcoxon signed-rank test, first applied to classifier comparison by Demšar [25], is a nonparametric hypothesis test that enables paired comparison of two models without assuming normality of score distributions. Bonferroni correction, originally formulated by Dunn [26] from Bonferroni’s inequality, controls the family-wise error rate in multiple simultaneous comparisons. Together, these two procedures provide robust statistical validation of XAI method differences across architectures. Intersection over Union (IoU) against manually annotated lung segmentation masks has been used to assess spatial localization quality [27], yet no study has combined all three metrics across three architecture families with full statistical validation.

This study addresses these gaps with three specific aims. First, we evaluate whether the proposed method — a compact, parameter-efficient architecture (EfficientNet-B0) — can outperform larger CNN baselines on pneumonia detection under identical training conditions. Second, we determine whether the highest-accuracy model also produces the best XAI explanations, a relationship that is assumed but rarely tested empirically. Third, we investigate whether architecture-specific properties create systematic differences in XAI method behavior, including potential failure modes. To our knowledge, this is the first study to conduct a three-family CNN comparison (residual, dense, and compound-scaled) for XAI quality in chest X-ray classification, evaluated across three quantitative metrics with full pairwise statistical validation over a publicly available dataset with lung segmentation masks.

## 2. MATERIALS AND METHODS

### 2.1 Datasets

Two publicly available datasets were used in this study. The Kaggle Chest X-Ray Images dataset [15] comprises 5,840 frontal chest X-ray images from pediatric patients (pneumonia: 4,265; normal: 1,575), reflecting an approximately 2:1 class imbalance. Images were partitioned into training (70%), validation (15%), and test (15%) splits. For XAI localization evaluation, lung segmentation masks from the publicly available Montgomery County chest X-ray dataset [14] were used exclusively to provide ground-truth anatomical regions (n = 138 images). This dataset represents one of the standard benchmark collections with manually annotated lung segmentation masks in the chest X-ray community, and has been consistently used as the reference for spatial localization evaluation in medical imaging studies [16]. Lung anatomy is sufficiently consistent across patient populations to serve as a valid spatial localization proxy for chest X-ray classification tasks. Both datasets used in this study are publicly available and fully anonymized. No patient-identifiable information was accessed, and no new data were collected. Institutional ethics approval was therefore not required for this study.

### 2.2 Data Preprocessing and Augmentation

All images were resized to 224×224 pixels prior to model input. On-the-fly data augmentation was applied during training, including random rotations up to 15°, width and height shifts up to 10%, zoom up to 10%, horizontal flipping, and reflect-mode border fill. A critical preprocessing distinction applies to EfficientNet-B0: the architecture’s built-in preprocessing function was applied rather than standard [4].

**DenseNet121** employs dense connectivity in which each layer receives feature maps from all preceding layers (7.98M parameters, 74.7% Top-1 ImageNet accuracy), promoting feature reuse and gradient flow across depth [5].

**EfficientNet-B0** (the proposed method) uses compound scaling to jointly optimize network depth, width, and resolution (5.3M parameters, 77.1% Top-1 ImageNet accuracy). Its MBConv blocks with squeeze-and-excitation modules produce spatially focused feature maps, making it the most parameter-efficient architecture in this comparison [6]. Table 1 summarizes the three architectures.

**Table 1.**
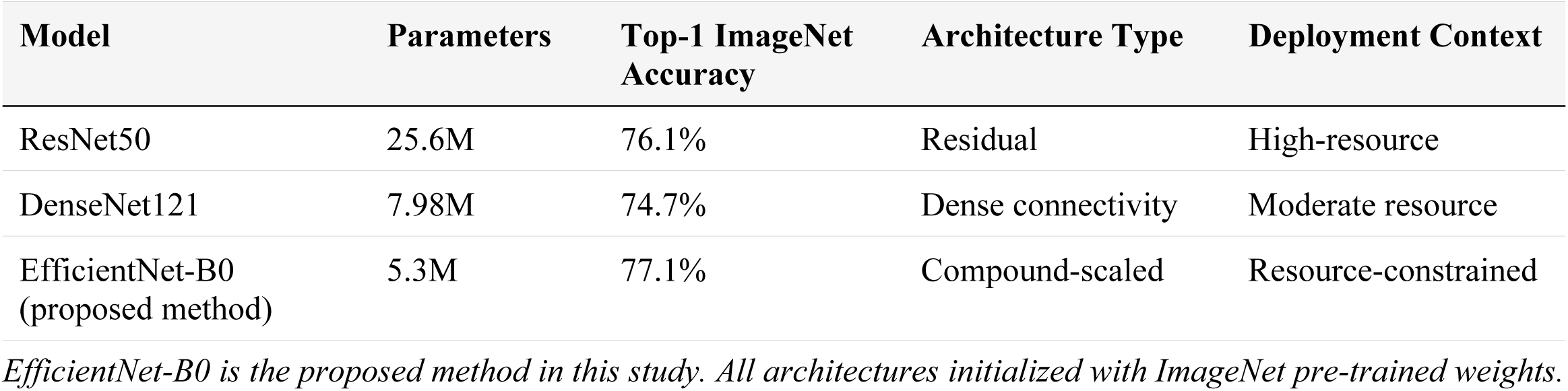
Summary of CNN architectures evaluated in this study.

| Model | Parameters | Top-1 ImageNet Accuracy | Architecture Type | Deployment Context |
| --- | --- | --- | --- | --- |
| ResNet50 | 25.6M | 76.1% | Residual | High-resource |
| DenseNet121 | 7.98M | 74.7% | Dense connectivity | Moderate resource |
| EfficientNet-B0 (proposed method) | 5.3M | 77.1% | Compound-scaled | Resource-constrained |

*EfficientNet-B0 is the proposed method in this study. All architectures initialized with ImageNet pre-trained weights.*

### 2.4 Training Protocol

A two-phase transfer learning protocol was applied identically across all three architectures. In Phase 1, backbone weights were frozen and only the classification head was trained for up to 50 epochs using the Adam optimizer with a learning rate of 0.0001. In Phase 2, the top 30 backbone layers were unfrozen for fine-tuning with a reduced learning rate of 0.00001 for up to 20 additional epochs. Both phases employed early stopping with a patience of 10 epochs, ReduceLROnPlateau scheduling (factor = 0.2, patience = 5 epochs), and ModelCheckpoint saving on minimum validation loss. Batch size was 32 for ResNet50 and DenseNet121, and 16 for EfficientNet-B0. Class imbalance was addressed for EfficientNet-B0 via scikit-learn’s compute_class_weight function with the’balanced’ setting; without this correction, the model converged to predicting only the pneumonia class (62% accuracy, 0% recall for normal), consistent with known sensitivity of compound-scaled architectures to class imbalance in binary medical classification tasks.

### 2.5 XAI Methods

Three gradient-based and activation-based XAI visualization methods were implemented and applied to the final convolutional layer of each architecture.

**Grad-CAM** computes the gradient of the binary classification score with respect to the final convolutional layer feature maps, applies global average pooling to obtain per-channel weights, and produces a weighted, ReLU-activated linear combination of the feature maps [8].

**Grad-CAM++** extends Grad-CAM by replacing global average-pooled gradients with pixel-wise second-and third-order partial derivatives as channel weighting coefficients, improving spatial localization for diffuse or multi-instance activation patterns [9].

**Score-CAM** eliminates gradient dependency by masking the input image with each upsampled activation channel individually and using the resulting change in model confidence as the channel weight. The final heatmap is a weighted sum of activation channels [10]. For EfficientNet-B0, all Score-CAM intermediate masked images and all Insertion/Deletion pixel-modified images were passed through EfficientNet’s built-in preprocessing function prior to model inference; omitting this step produces invalid metric values.

### 2.6 Score-CAM Channel Selection

Score-CAM requires selecting a channel budget due to the computational cost of per-channel forward passes. Channel budgets of 20, 50, 100, and all available channels were evaluated on a subset of segmentation-mask images for each architecture. Based on this analysis (see Results), a budget of 20 channels was selected for ResNet50, which exhibited monotonically decreasing IoU with increasing channel count. A budget of 100 channels was selected for both DenseNet121 and EfficientNet-B0, where IoU plateaued between 50 and 100 channels.

### 2.7 Quantitative Evaluation Metrics

Three quantitative metrics were used to evaluate XAI explanation quality across all nine model– method combinations.

**Intersection over Union (IoU)** measures the spatial overlap between the binarized XAI heatmap and the ground-truth lung segmentation mask. Heatmaps were binarized at five thresholds (0.3, 0.4, 0.5, 0.6, 0.7) and IoU was computed at each threshold and averaged to produce a threshold-robust mean IoU. Higher values indicate better anatomical localization (↑).

**Insertion score** measures XAI faithfulness by starting from a Gaussian-blurred image and progressively revealing pixels in order of their assigned importance, computing the area under the resulting model confidence curve. Higher values indicate that the most important pixels drive model confidence (↑).

**Deletion score** is the complementary measure, starting from the original image and progressively removing pixels in order of importance, computing the area under the resulting confidence curve. Lower values indicate that the most important pixels, when removed, most strongly reduce model confidence (↓). All three metrics were computed over all available segmentation-mask images for each of the nine model–method combinations (n = 138 images).

### 2.8 Statistical Analysis

Pairwise Wilcoxon signed-rank tests were conducted for all three XAI method pairs (Grad-CAM vs. Grad-CAM++, Grad-CAM vs. Score-CAM, Grad-CAM++ vs. Score-CAM) within each model–metric combination, yielding 27 tests in total. Bonferroni correction was applied within each model–metric group (three pairs per group), giving a corrected significance threshold of α = 0.0167. Effect size was quantified using rank-biserial correlation (r), with |r| > 0.5 indicating large practical significance. All statistical analyses were conducted in Python using the SciPy library.

## 3. RESULTS

### 3.1 Classification Performance

All three models achieved strong classification performance on the held-out test set (Table 2). EfficientNet-B0 (the proposed method) outperformed both baselines across all metrics while using the fewest parameters: 0.90 accuracy and AUC 0.97, compared to DenseNet121 (0.88, AUC 0.96) and ResNet50 (0.84, AUC 0.92). EfficientNet-B0 also achieved the lowest false negative count on the test set (9 false negatives), matching DenseNet121 and outperforming ResNet50 (21 false negatives) — a clinically relevant distinction given that missed pneumonia diagnoses carry greater risk than false positives. Training history curves for all three models are shown in Fig 1; all models converged stably. The EfficientNet-B0 training history shows a marked improvement in validation accuracy following Phase 2 fine-tuning, consistently exceeding the DenseNet121 baseline of 88%. Confusion matrices for the test set are presented in Fig 2.

**Fig 1.**
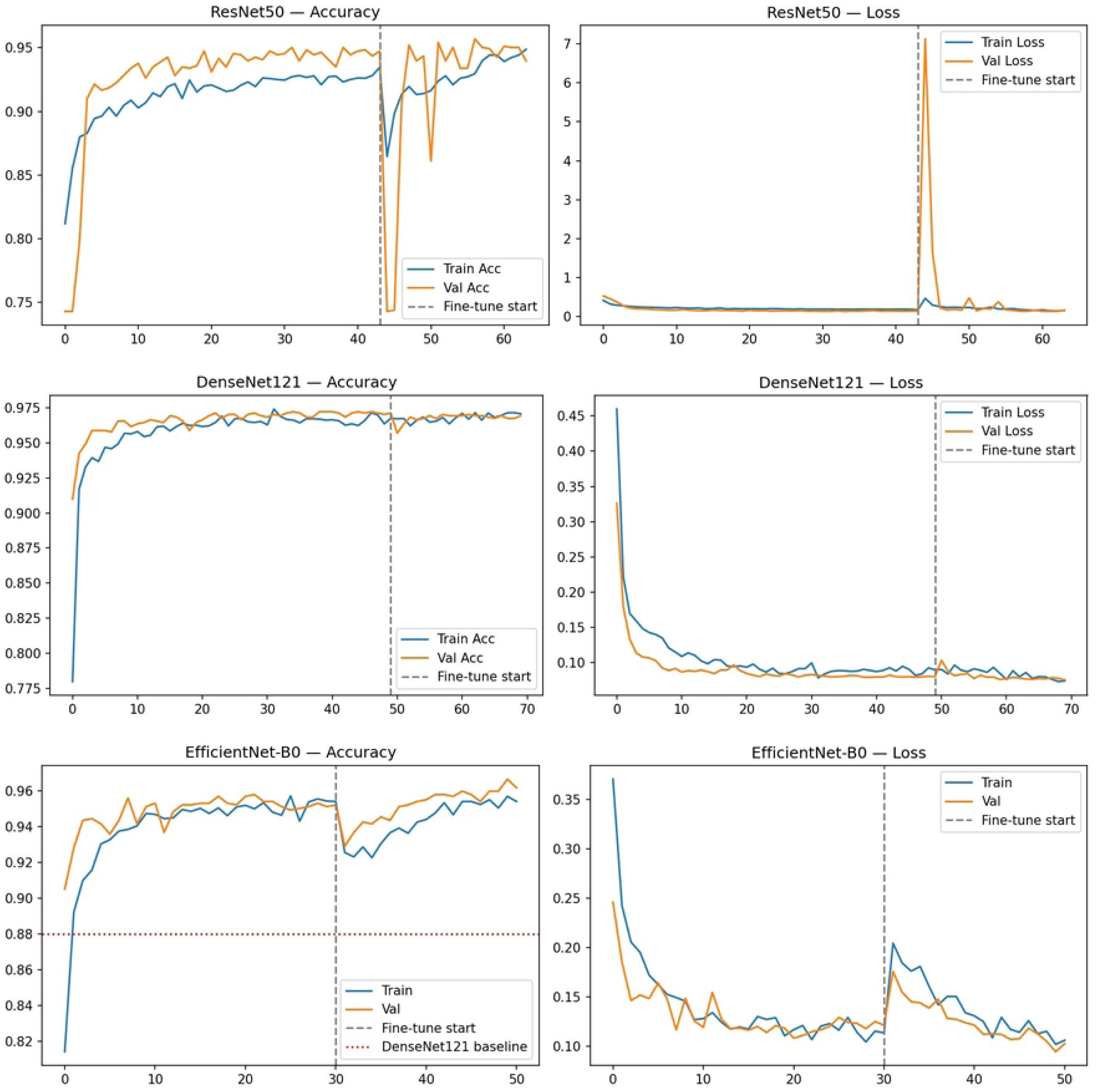
Training accuracy and loss curves for all three models. Phase 1 corresponds to frozen backbone training; Phase 2 (indicated by the dashed vertical line) corresponds to fine-tuning of the top 30 backbone layers. Panels show ResNet50 (top), DenseNet121 (middle), and EfficientNet-B0 / proposed method (bottom). The horizontal dotted line in the bottom panel indicates the DenseNet121 validation accuracy baseline of 88%.

**Fig 2.**
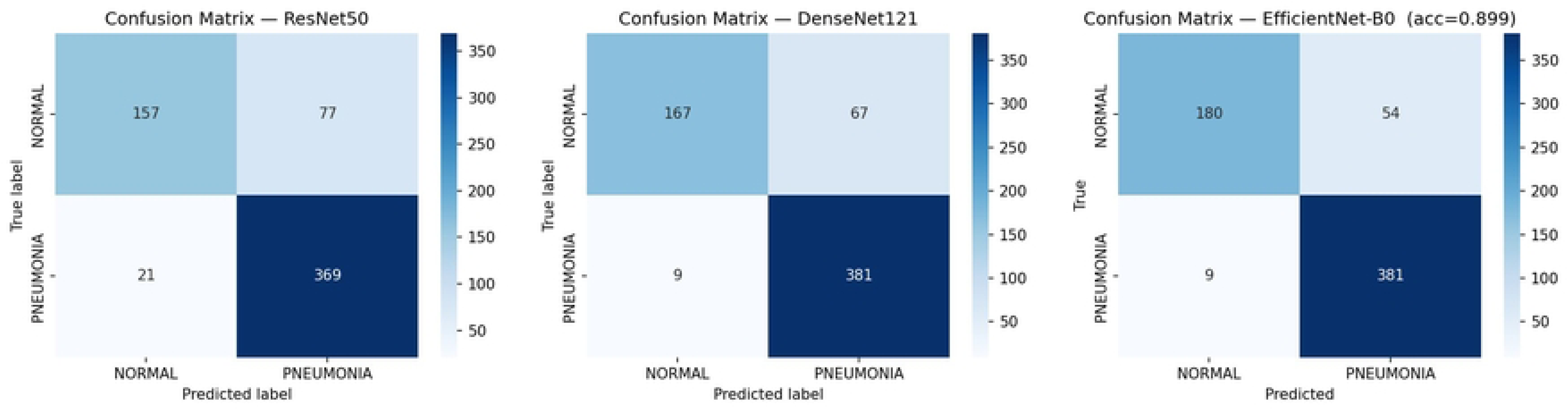
Confusion matrices on the held-out test set for ResNet50 (left), DenseNet121 (center), and EfficientNet-B0 / proposed method (right). Rows indicate true class; columns indicate predicted class. Values represent number of images.

**Table 2.** Classification performance on the chest X-ray test set.

| Model | Accuracy | AUC | F1-Score | Average Precision | Parameters |
| --- | --- | --- | --- | --- | --- |
| ResNet50 | 0.84 | 0.921 | 0.84 | 0.947 | 25.6M |
| DenseNet121 | 0.88 | 0.964 | 0.87 | 0.976 | 7.98M |
| EfficientNet-B0 (proposed method) | 0.90 | 0.973 | 0.90 | 0.984 | 5.3M |

All models trained under an identical two-phase transfer learning protocol on the Kaggle chest X-ray dataset ( $n = 5,863$ ). AUC = area under the receiver operating characteristic curve.

### 3.2 Qualitative XAI Analysis

Representative XAI heatmaps for a normal and a pneumonia chest X-ray are shown in Fig 3 and Fig 4, respectively. For the normal image (Fig 3), DenseNet121 with Score-CAM produces the most spatially distributed coverage of the lung fields, consistent with its superior mean IoU reported below. The proposed method (EfficientNet-B0) with Score-CAM shows broad bilateral lung activation, while ResNet50 with Grad-CAM produces more concentrated activations that are anatomically less consistent across images. For the pneumonia case (Fig 4), DenseNet121 and the proposed method correctly focus on the right lower lobe consolidation region. ResNet50 with Score-CAM produces a sparse heatmap localized to a small region of the lung field, qualitatively consistent with the quantitative Score-CAM failure mode reported in the channel sensitivity analysis below.

**Fig 3.**
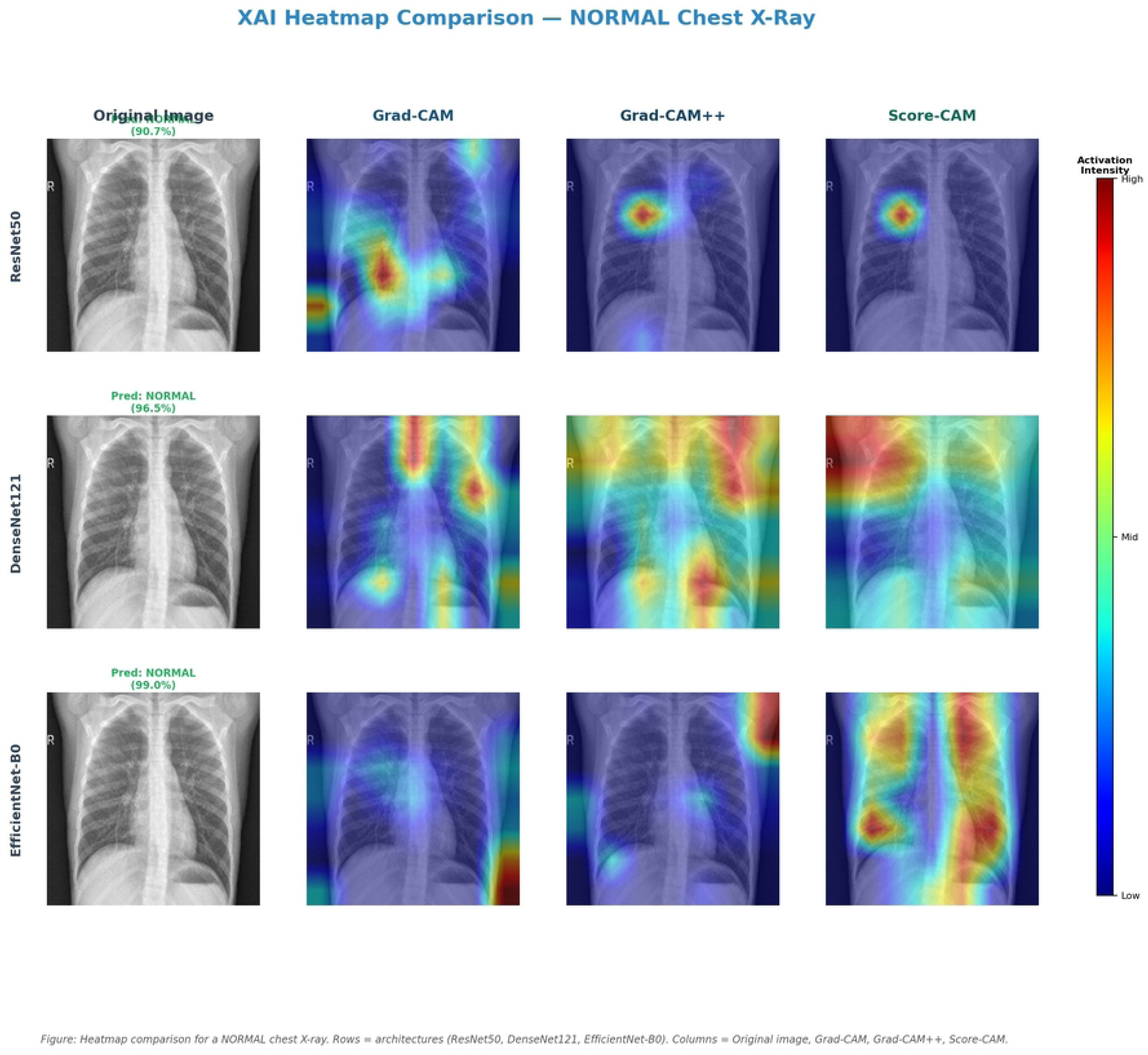
XAI heatmap comparison for a representative normal chest X-ray. Rows correspond to architectures (ResNet50, DenseNet121, EfficientNet-B0 / proposed method). Columns correspond to the original image, Grad-CAM, Grad-CAM++, and Score-CAM overlays. Warmer colors indicate higher activation.

**Fig 4.**
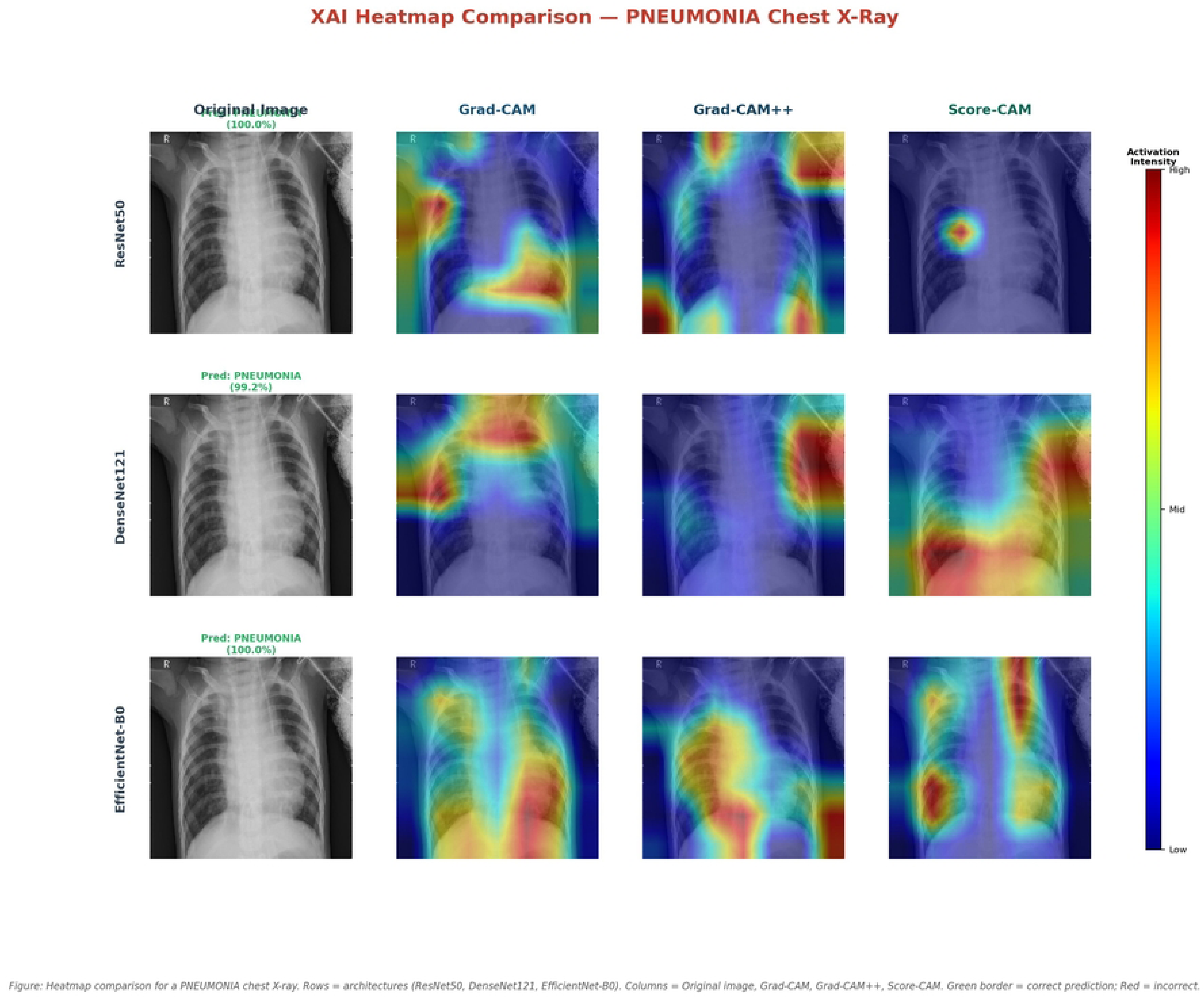
XAI heatmap comparison for a representative pneumonia chest X-ray. Rows correspond to architectures (ResNet50, DenseNet121, EfficientNet-B0 / proposed method). Columns correspond to the original image, Grad-CAM, Grad-CAM++, and Score-CAM overlays. Warmer colors indicate higher activation.

### 3.3 Quantitative XAI Evaluation

Table 3 presents mean IoU, Insertion score, and Deletion score for all nine model–method combinations (n = 138 images). Fig 5 provides a visual summary across all combinations.

**Fig 5.**
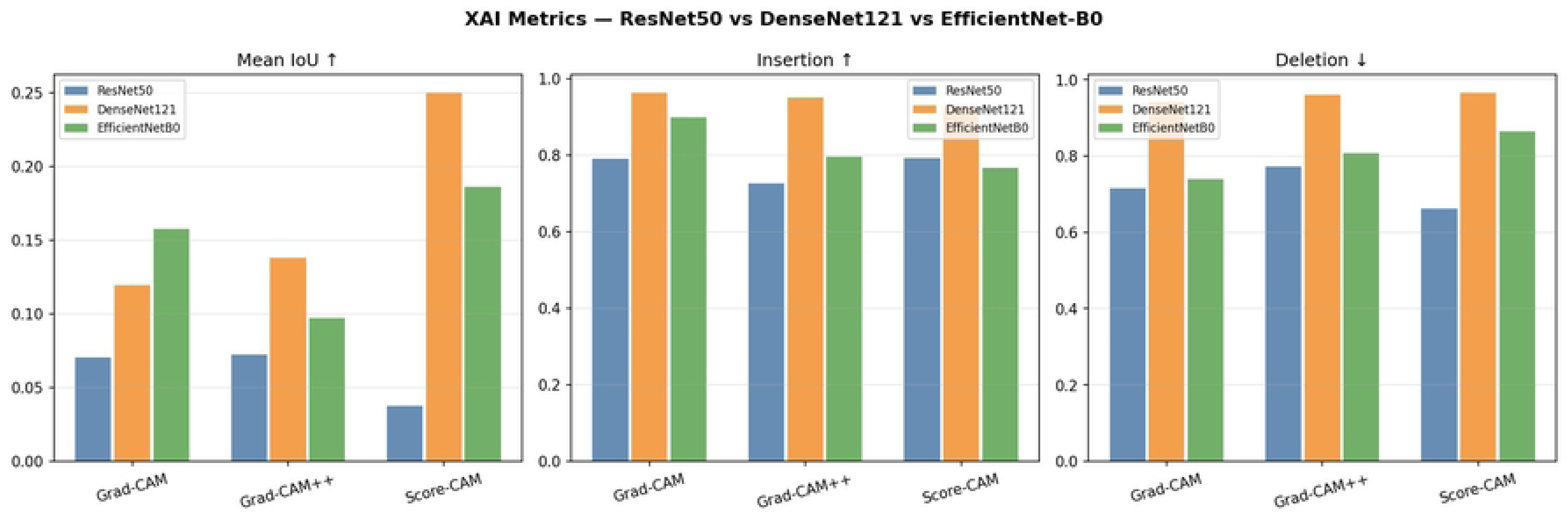
Bar chart of mean IoU, Insertion score, and Deletion score across all nine model–method combinations. Error bars represent one standard deviation. For Deletion score, lower values indicate better performance.

**Table 3.**
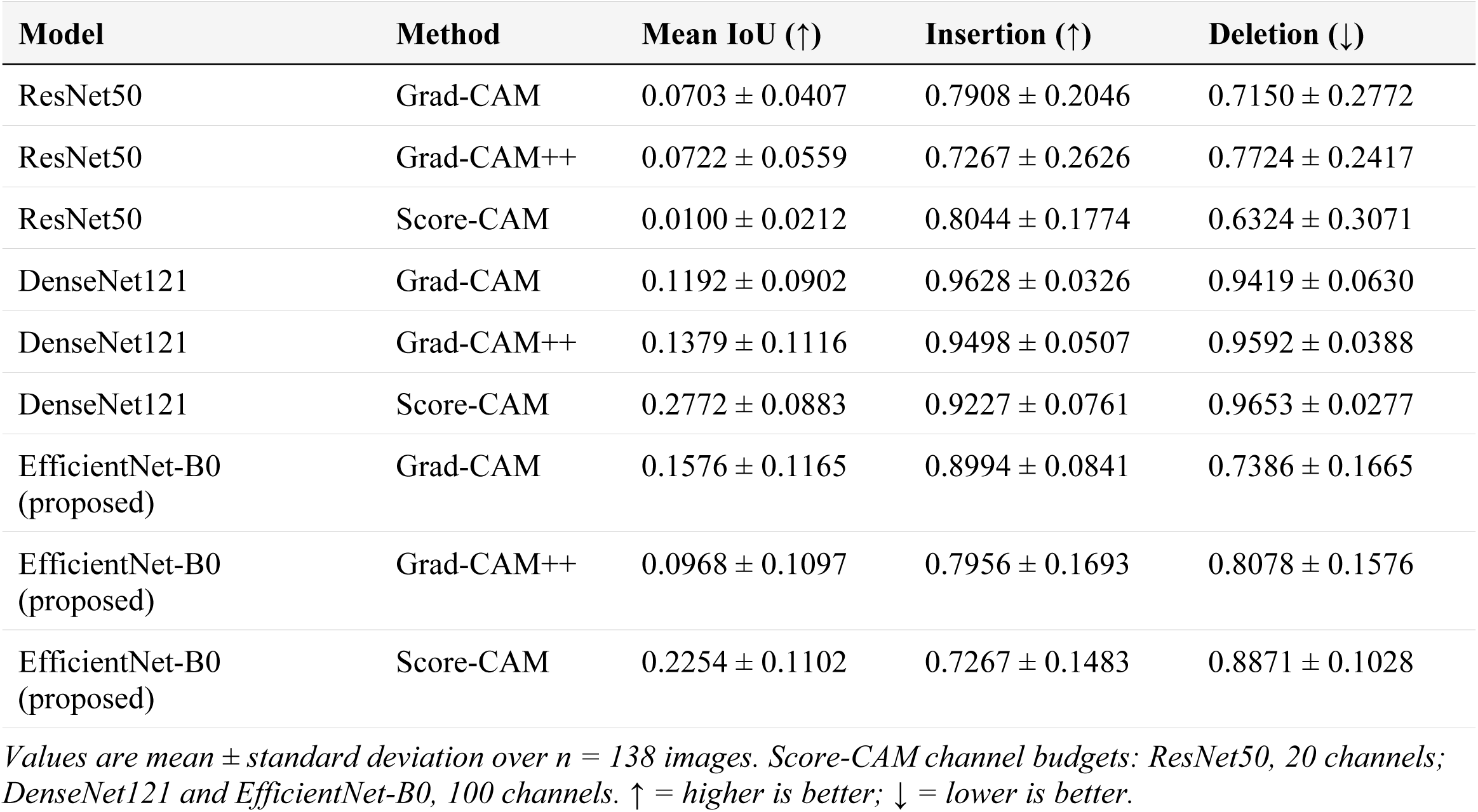
Quantitative XAI evaluation results for all nine model–method combinations.

| Model | Method | Mean IoU (↑) | Insertion (↑) | Deletion (↓) |
| --- | --- | --- | --- | --- |
| ResNet50 | Grad-CAM | $0.0703 \pm 0.0407$ | $0.7908 \pm 0.2046$ | $0.7150 \pm 0.2772$ |
| ResNet50 | Grad-CAM++ | $0.0722 \pm 0.0559$ | $0.7267 \pm 0.2626$ | $0.7724 \pm 0.2417$ |
| ResNet50 | Score-CAM | $0.0100 \pm 0.0212$ | $0.8044 \pm 0.1774$ | $0.6324 \pm 0.3071$ |
| DenseNet121 | Grad-CAM | $0.1192 \pm 0.0902$ | $0.9628 \pm 0.0326$ | $0.9419 \pm 0.0630$ |
| DenseNet121 | Grad-CAM++ | $0.1379 \pm 0.1116$ | $0.9498 \pm 0.0507$ | $0.9592 \pm 0.0388$ |
| DenseNet121 | Score-CAM | $0.2772 \pm 0.0883$ | $0.9227 \pm 0.0761$ | $0.9653 \pm 0.0277$ |
| EfficientNet-B0 (proposed) | Grad-CAM | $0.1576 \pm 0.1165$ | $0.8994 \pm 0.0841$ | $0.7386 \pm 0.1665$ |
| EfficientNet-B0 (proposed) | Grad-CAM++ | $0.0968 \pm 0.1097$ | $0.7956 \pm 0.1693$ | $0.8078 \pm 0.1576$ |
| EfficientNet-B0 (proposed) | Score-CAM | $0.2254 \pm 0.1102$ | $0.7267 \pm 0.1483$ | $0.8871 \pm 0.1028$ |

Values are mean ± standard deviation over $n = 138$ images. Score-CAM channel budgets: ResNet50, 20 channels; DenseNet121 and EfficientNet-B0, 100 channels. ↑ = higher is better; ↓ = lower is better.

DenseNet121 with Score-CAM achieved the highest mean IoU (0.2772 ± 0.0883), indicating the best spatial alignment with ground-truth lung masks. EfficientNet-B0 with Score-CAM ranked second on IoU (0.2254 ± 0.1102), followed by EfficientNet-B0 with Grad-CAM (0.1576 ± 0.1165). All ResNet50 combinations produced substantially lower IoU values, with ResNet50 Score-CAM achieving the lowest mean IoU of any combination (0.0100 ± 0.0212).

For Insertion score, DenseNet121 with Grad-CAM achieved the highest value (0.9628 ± 0.0326), indicating strong faithfulness of pixel importance rankings. DenseNet121 consistently produced the highest Insertion scores across all three XAI methods (0.9227–0.9628) with the lowest standard deviations (0.0277–0.0761). For Deletion score, ResNet50 with Score-CAM achieved the lowest mean (0.6324 ± 0.3071); however, the large standard deviation indicates high cross-image variability, limiting the reliability of this result.

### 3.4 Statistical Significance of XAI Method Differences

Of 27 pairwise Wilcoxon signed-rank tests, 24 were statistically significant after Bonferroni correction (Fig 6). Three pairs were non-significant: ResNet50 Grad-CAM vs. Grad-CAM++ on IoU (both near-zero localization, p = 1.0); ResNet50 Grad-CAM vs. Score-CAM on Insertion (overlapping confidence distributions, p = 1.0); and DenseNet121 Grad-CAM vs. Grad-CAM++ on IoU (p = 0.18). These non-significant results are interpretable given the underlying metric distributions. Large effect sizes (|r| > 0.5) were observed in 11 of 27 pairs, with the largest being EfficientNet-B0 Grad-CAM vs. Score-CAM on Insertion score (r = −0.805).

**Fig 6.**
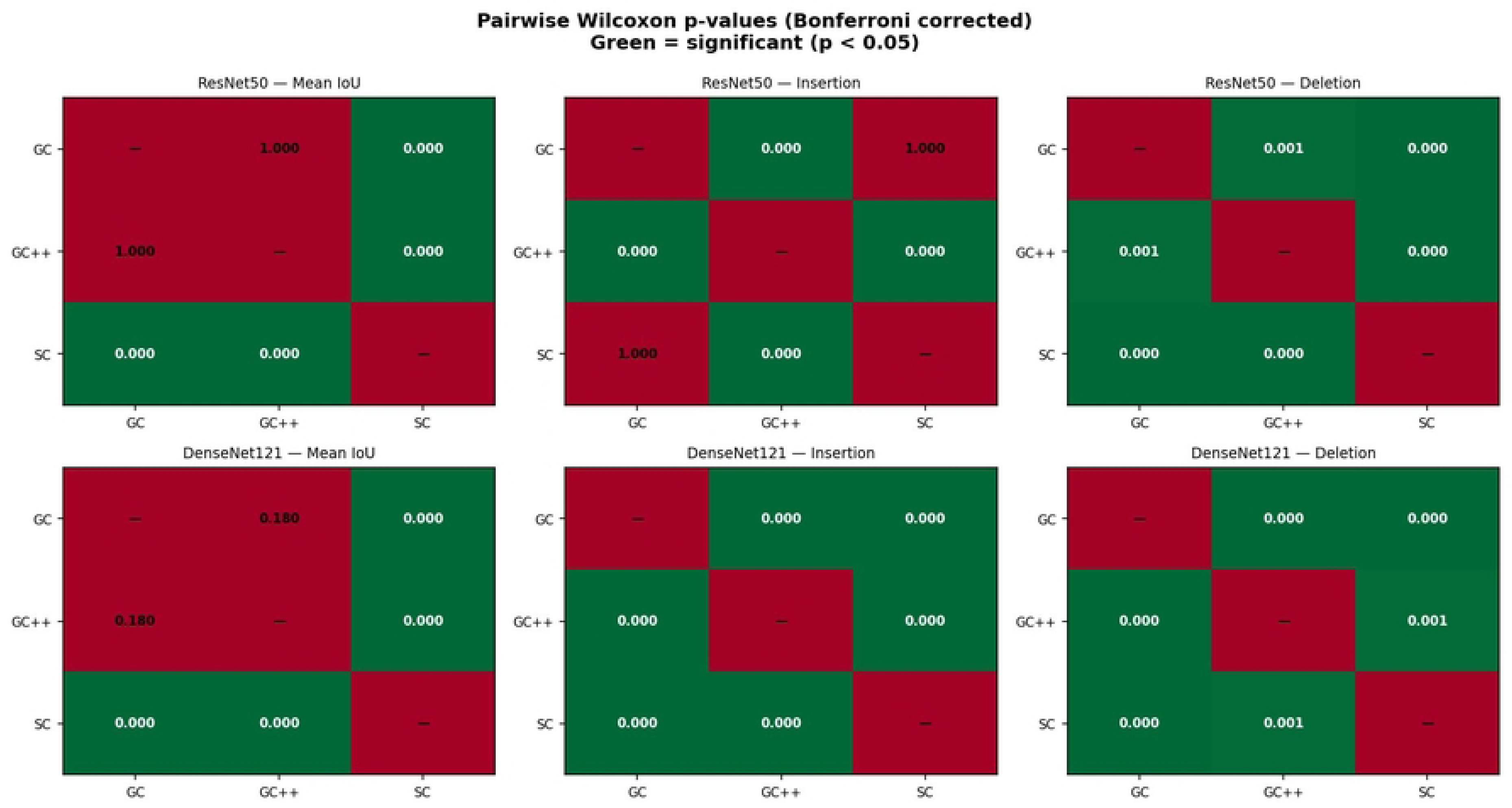
Pairwise Wilcoxon signed-rank test p-values with Bonferroni correction across all model–metric combinations (n = 138). Green cells indicate statistically significant differences (p < 0.05 after correction); red cells indicate non-significant pairs. GC = Grad-CAM; GC++ = Grad-CAM++; SC = Score-CAM.

### 3.5 Per-Threshold IoU Analysis

Table 4 presents IoU at each individual binarization threshold. DenseNet121 with Score-CAM maintained the highest IoU across all five thresholds, reflecting spatially precise heatmaps that remain well-localized at stricter binarization levels. EfficientNet-B0 with Score-CAM achieved the highest single-threshold IoU at t = 0.3 (0.3526), exceeding DenseNet121 Score-CAM (0.3113) at that threshold, suggesting broad but anatomically relevant spatial coverage. IoU values declined monotonically with increasing threshold for all combinations, consistent with progressive heatmap binarization reducing overlap area.

**Table 4.** IoU scores at individual binarization thresholds for all nine model–method combinations.

| Model | Method | IoU@0.3 | IoU@0.4 | IoU@0.5 | IoU@0.6 | IoU@0.7 |
| --- | --- | --- | --- | --- | --- | --- |
| ResNet50 | Grad-CAM | 0.1150 | 0.0911 | 0.0685 | 0.0482 | 0.0286 |
| ResNet50 | Grad-CAM++ | 0.1308 | 0.0976 | 0.0687 | 0.0417 | 0.0219 |
| ResNet50 | Score-CAM | 0.0187 | 0.0136 | 0.0091 | 0.0056 | 0.0031 |
| DenseNet121 | Grad-CAM | 0.1771 | 0.1483 | 0.1180 | 0.0904 | 0.0619 |
| DenseNet121 | Grad-CAM++ | 0.1956 | 0.1677 | 0.1402 | 0.1099 | 0.0760 |
| DenseNet121 | Score-CAM | 0.3113 | 0.3092 | 0.2945 | 0.2637 | 0.2072 |
| EfficientNet-B0 (proposed) | Grad-CAM | 0.2556 | 0.2080 | 0.1549 | 0.1060 | 0.0636 |
| EfficientNet-B0 (proposed) | Grad-CAM++ | 0.1562 | 0.1274 | 0.0961 | 0.0657 | 0.0387 |
| EfficientNet-B0<br>(proposed) | Score-CAM | 0.3526* | 0.2955 | 0.2287 | 0.1569 | 0.0934 |

\*Highest single-threshold IoU across all nine combinations at $t = 0.3$ .

### 3.6 Score-CAM Channel Sensitivity Analysis

Table 5 presents the Score-CAM channel budget sensitivity analysis. ResNet50 exhibited monotonically decreasing IoU with increasing channel count, collapsing to zero at the full 2048-channel budget, confirming non-discriminative channel dominance as the mechanism underlying Score-CAM failure for this architecture. DenseNet121 and EfficientNet-B0 both showed IoU plateaus between 50 and 100 channels, with marginal gains beyond 100 channels at substantially higher computational cost. The full-channel budget for EfficientNet-B0 (320 channels) produced only a marginal IoU improvement (0.2724 vs. 0.2526 at 100 channels) at 14× greater runtime, indicating that 100 channels represents the optimal efficiency-accuracy trade-off for these architectures.

**Table 5.** Score-CAM channel budget sensitivity analysis.

| Model | Channels | Mean IoU ± SD | Runtime (s) | Selected |
| --- | --- | --- | --- | --- |
| ResNet50 | 20 | 0.0398 ± 0.0387 | 85 | Yes |
| ResNet50 | 50 | 0.0243 ± 0.0280 | 212 | No |
| ResNet50 | 100 | 0.0069 ± 0.0173 | 440 | No |
| ResNet50 | 2048 (all) | 0.0000 ± 0.0000 | 8496 | No |
| DenseNet121 | 20 | 0.2168 ± 0.0925 | 132 | No |
| DenseNet121 | 50 | 0.2555 ± 0.0977 | 207 | No |
| DenseNet121 | 100 | 0.2555 ± 0.0977 | 220 | Yes |
| EfficientNet-B0 | 20 | 0.1956 ± 0.1451 | 91 | No |
| EfficientNet-B0 | 50 | 0.2660 ± 0.1276 | 220 | No |
| EfficientNet-B0 | 100 | 0.2526 ± 0.1173 | 442 | Yes |
| EfficientNet-B0 | 320 (all) | 0.2724 ± 0.1293 | 6295 | No |

*Selected channel budgets are those that maximized mean IoU within a practical runtime constraint.*

## 4. DISCUSSION

### 4.1 Parameter Efficiency and Classification Performance

The central practical finding of this study is that the proposed method (EfficientNet-B0) achieves the highest classification accuracy among the three architectures evaluated while using the fewest parameters. The 4.8× parameter efficiency advantage over ResNet50 is attributable to compound scaling: jointly optimizing network depth, width, and resolution via a neural architecture search-derived coefficient produces a more favorable inductive bias for binary chest X-ray classification than either residual skip connections or dense feature reuse alone. This result is consistent with EfficientNet-B0’s superior Top-1 ImageNet accuracy (77.1%) relative to ResNet50 (76.1%) and DenseNet121 (74.7%), suggesting that the compound-scaling advantage generalizes from natural image classification to medical imaging tasks under transfer learning conditions.

A practically important implementation finding concerns class imbalance handling. Without explicit class weighting via scikit-learn’s compute_class_weight function, EfficientNet-B0 converged to predicting only the pneumonia class (62% accuracy, 0% recall for normal), a behavior not observed for ResNet50 or DenseNet121 under identical conditions. This suggests that compound-scaled architectures may be more sensitive to class imbalance in binary medical classification tasks, and that explicit class weighting should be treated as a standard implementation requirement when training EfficientNet variants on imbalanced medical datasets.

For clinical deployment contexts where computational resources are constrained — including mobile screening units, edge devices, and high-throughput cloud APIs — the proposed method offers a favorable combination of superior accuracy and substantially reduced memory and inference cost relative to both baselines. The 9 false negatives recorded by EfficientNet-B0 on the test set, matching DenseNet121 and substantially fewer than ResNet50 (21 false negatives), further supports its suitability for screening applications where missed diagnoses carry greater clinical risk than false positives.

### 4.2 Localization Quality and the Score-CAM Failure Mode

DenseNet121 with Score-CAM achieved the highest mean IoU (0.2772 ± 0.0883), indicating that dense connectivity produces activation patterns that are most spatially aligned with ground-truth lung anatomy. This result is consistent with the architectural property of DenseNet: by aggregating feature maps from all preceding layers, dense connectivity encourages the network to retain spatially precise low-level features alongside high-level semantic representations, producing more anatomically coherent activations at the final convolutional layer.

The most architecturally informative finding in this study is the Score-CAM failure mode specific to ResNet50. Score-CAM produced the worst IoU for ResNet50 (0.0100 ± 0.0212) — lower than Grad-CAM and Grad-CAM++ by an order of magnitude — yet produced the best or second-best IoU for DenseNet121 and EfficientNet-B0. The channel sensitivity analysis reveals the underlying mechanism: ResNet50’s 2048-channel final layer is dominated by non-discriminative channels that receive near-zero forward-pass confidence scores when used as image masks. As more channels are included in the Score-CAM weighted sum, the heatmap collapses monotonically toward zero, eventually producing a blank activation map at the full 2048-channel budget. Grad-CAM’s gradient-based weighting mechanism is robust to this effect because the gradient signal inherently suppresses inactive channels, regardless of their count. This failure mode would not be detectable through qualitative heatmap inspection alone, underscoring the necessity of quantitative evaluation when selecting XAI methods for a given architecture. Practitioners using Score-CAM with ResNet-family architectures should apply aggressive channel selection or consider gradient-based alternatives.

The proposed method (EfficientNet-B0) with Grad-CAM ranked second overall on mean IoU (0.1576 ± 0.1165), and EfficientNet-B0 with Score-CAM ranked third (0.2254 ± 0.1102). The higher IoU of Score-CAM over Grad-CAM for EfficientNet-B0 — the reverse of the ResNet50 pattern — reflects the more moderate channel count of EfficientNet-B0’s final layer (320 channels vs. 2048), which does not trigger the same collapse mechanism.

### 4.3 Faithfulness and Architectural Encoding Properties

DenseNet121 consistently achieved the highest Insertion scores across all three XAI methods (0.9227–0.9628), with the lowest standard deviations (0.0277–0.0761). This stability reflects the redundancy of dense feature encoding: because each layer aggregates all preceding feature maps, the most important pixels identified by any XAI method are likely to be genuinely discriminative, producing reliable Insertion curves across images. EfficientNet-B0’s intermediate Insertion scores (0.7267–0.8994) reflect the more distributed spatial encoding of MBConv depthwise separable convolutions, which decompose spatial and channel-wise feature learning. This is an architectural property rather than a failure of the XAI methods, and it suggests that faithfulness metrics may be systematically influenced by architectural encoding strategy independently of classification performance.

ResNet50 with Score-CAM achieved the lowest Deletion score (0.6324 ± 0.3071), which might superficially appear to indicate the best pixel faithfulness. However, the large standard deviation indicates high cross-image variability, and this result is mechanistically explained by the Score-CAM heatmap collapse: when the heatmap is effectively blank, the pixels removed first during Deletion are essentially random, producing an artificially favorable Deletion curve. This result therefore does not reflect genuine XAI quality and should not be interpreted as such.

### 4.4 Accuracy and Interpretability as Independent Properties

The empirical finding that the highest-accuracy model does not produce the best XAI explanations is the central scientific contribution of this study. The proposed method (EfficientNet-B0) achieved 90% classification accuracy but did not lead on any XAI metric. DenseNet121, with 88% accuracy, produced the best spatial localization and highest faithfulness scores. This dissociation was statistically confirmed across 24 of 27 pairwise comparisons, with 11 large effect sizes (|r| > 0.5), indicating that the accuracy-interpretability gap is not a noise artifact but a genuine and reproducible architectural property. These findings have direct implications for clinical deployment decisions: selecting a model on the basis of classification accuracy alone is insufficient if XAI explanation quality is a deployment requirement.

Based on these results, the following evidence-based recommendations can be made. The proposed method (EfficientNet-B0) is recommended for accuracy-critical or resource-constrained settings, where its superior classification performance (90% accuracy, AUC 0.973) and reduced computational footprint (5.3M parameters) outweigh its intermediate XAI performance. DenseNet121 with Score-CAM is recommended for interpretability-critical settings where spatial localization quality is the primary requirement, offering the best anatomical alignment with ground-truth lung regions (mean IoU = 0.2772). DenseNet121 with Grad-CAM is recommended for faithfulness-critical settings where pixel importance ranking consistency is prioritized, offering the highest Insertion score (0.9628 ± 0.0326) and lowest standard deviation across all combinations. Score-CAM should be avoided with ResNet50 due to the channel collapse failure mode identified in this study; Grad-CAM is the preferred XAI method for ResNet-family architectures.

### 4.5 Limitations

Several limitations of this study should be acknowledged. First, the IoU evaluation used adult lung segmentation masks from the Montgomery County dataset, while the Kaggle chest X-ray dataset comprises pediatric patients. Differences in lung geometry between pediatric and adult populations may affect IoU values systematically, and validation using pediatric-specific segmentation masks is recommended. Second, only EfficientNet-B0 was evaluated among the EfficientNet family; larger variants (B1–B7) and the more recent EfficientNetV2 may exhibit different XAI properties and should be investigated in future work with appropriate GPU resources. Third, the Score-CAM channel budget was calibrated on a subset of 20 images, and optimal budgets may differ across datasets or clinical imaging protocols. Fourth, no clinician-centered user study was conducted to assess whether quantitatively superior heatmaps improve diagnostic confidence or decision accuracy in practice; such a study would be an important complement to the quantitative evaluation presented here. Fifth, generalizability to multi-class chest X-ray classification tasks, such as those in the NIH ChestX-ray14 dataset [31], was not evaluated.

### 4.6 Conclusions

This study provides a systematic, statistically validated comparison of three XAI methods across three CNN architecture families for pneumonia detection from chest X-ray images. The proposed method (EfficientNet-B0) achieved the highest classification accuracy (90%, AUC 0.973) using the fewest parameters (5.3M), establishing it as the preferred architecture for resource-constrained clinical deployment. However, the proposed method did not produce the best XAI explanations, confirming that accuracy and interpretability must be evaluated and optimized independently. DenseNet121 with Score-CAM achieved the best spatial localization (mean IoU = 0.2772 ± 0.0883), and DenseNet121 with Grad-CAM achieved the highest faithfulness (Insertion = 0.9628 ± 0.0326). A novel Score-CAM failure mode was identified in ResNet50’s 2048-channel final layer, demonstrating that architecture-specific properties can cause systematic XAI method degradation that is undetectable through qualitative inspection alone. Statistical validation confirmed that 24 of 27 pairwise XAI method comparisons were significant after Bonferroni correction, with 11 large effect sizes. Future work should extend this evaluation to larger EfficientNet variants, Vision Transformers, and multi-class chest X-ray datasets, and should incorporate radiologist-centered user studies to assess the clinical utility of quantitatively superior heatmaps.

## Data Availability

No new data were generated by this study. The following existing data sources were used: the Kaggle Chest X-Ray Images (Pneumonia) dataset, available at https://www.kaggle.com/datasets/paultimothymooney/chest-xray-pneumonia and the Montgomery County chest X-ray collection, available from the National Library of Medicine at https://openi.nlm.nih.gov/imgs/collections/NLM-MontgomeryCXRSet.zip Both datasets are publicly available in full, and no additional data were generated in the course of this study.

https://www.kaggle.com/datasets/paultimothymooney/chest-xray-pneumonia

## ACKNOWLEDGMENTS

The authors would like to thank the Department of Computer Engineering, Faculty of Engineering and Technology, Shahid Ashrafi Esfahani University for providing the computational resources and academic environment that supported this research. The authors also gratefully acknowledge the open-access chest X-ray dataset made available through Kaggle by Paul Mooney and the Guangzhou Women and Children’s Medical Center, without which this work would not have been possible. No specific grant from any funding agency in the public, commercial, or not-for-profit sectors was received for this research.

## AUTHOR CONTRIBUTIONS

Benyamin Mahtabi: Conceptualization, Methodology, Software, Formal analysis, Investigation, Data curation, Writing – original draft, Visualization.

Ebrahim Nasr-Esfahani: Supervision, Writing – review & editing, Funding acquisition. Shokufeh Yaraghi: Supervision, Writing – review & editing, Funding acquisition.

## FUNDING

The authors received no specific funding for this work.

## COMPETING INTERESTS

The authors have declared that no competing interests exist.

## DATA AVAILABILITY STATEMENT

All data underlying the results presented in this study are publicly available. The chest X-ray images used for model training and evaluation are available from the Kaggle Chest X-Ray Images dataset (https://www.kaggle.com/datasets/paultimothymooney/chest-xray-pneumonia). The lung segmentation masks used for XAI localization evaluation are available from the Montgomery County chest X-ray collection via the National Library of Medicine (https://openi.nlm.nih.gov). No additional data were generated in this study.

## Notes

### Competing Interest Statement

The authors have declared no competing interest.

### Author Declarations

This study did not involve the collection of new data from human participants. All chest X-ray images and lung segmentation masks used were obtained from pre-existing, publicly available, and fully de-identified datasets (the Kaggle Chest X-Ray Images (Pneumonia) dataset and the Montgomery County chest X-ray collection via the National Library of Medicine). As no identifiable human participant data was accessed or generated, this study did not meet the threshold requiring IRB review or exemption determination at the authors' institution, Shahid Ashrafi Esfahani University.

